# Application of deep learning to classify skeletal growth phase on 3D radiographs

**DOI:** 10.1101/2023.03.08.23287008

**Authors:** Nazila Ameli, Manuel Lagravere, Hollis Lai

## Abstract

Cervical vertebral maturation (CVM) is widely used to evaluate growth potential in the field of orthodontics. The aim of this study is to develop an artificial intelligence (AI) algorithm to automatically predict the CVM stages in terms of growth phases using the cone-beam computed tomographic (CBCT) images. A total of 30,016 slices obtained from 56 patients with the age range of 7-16 years were included in the dataset. After cropping the region of interest (ROI), a convolutional neural network (CNN) was built to classify the slices based on the presence of a good vision of vertebrae for classification of the growth stages. The output was used to train another model capable of categorizing the slices into phases of growth, which were defined as Phase I (prepubertal, CVM stages 1 and 2), phase II (circumpubertal, CVM stage 3), and phase III (postpubertal, CVM stages 4, 5, and 6). After training the model, 88 unused images belonging to 3 phases were used to evaluate the performance of the model using multi-class classification metrics. The average classification accuracy of the first and second CNN-based deep learning models were 96.06% and 95.79%, respectively on the validation dataset. The multi-class classification metrics applied to the new testing dataset also showed an overall accuracy of 84% for predicting the growth phase. Moreover, phase I ranked the highest accuracy in terms of F1 score (87%), followed by phase II (83%), and phase III (80%) on new images. Our proposed models could automatically detect the C2-C4 vertebrae required for CVM staging and accurately classify slices into 3 growth phases without the need for annotating the shape and configuration of vertebrae. This will result in developing a fully automatic and less complex system with reasonable performance, comparable to expert practitioners.

**Author Summary:** The skeletal age of orthodontic patients is a critical factor in planning the proper orthodontic treatment. Thus, an accurate assessment of the growth stage can result in better treatment outcomes and reduced treatment time. Traditionally, 2-D cephalometric radiographs obtained during the orthodontic examination were used for estimating the skeletal age using the three cervical vertebrae. However, this method was subjective and prone to errors as different orthodontists could interpret the features differently. Moreover, 2-D images provide only limited information as they only capture two dimensions and involve superimpositions of neighbour structures. In the present study, machine learning models are applied to 3-D cephalometric images to predict the growth stage of patients by analyzing the shape and pattern of cervical vertebrae. This method has the potential to improve treatment outcomes and reduce the treatment time for orthodontic patients. Additionally, it can contribute to the development of more personalized treatment plans and advance our understanding of the growth and development of the craniofacial complex.

## Introduction

Understanding the growth and development process of children and adolescents is an important task in medicine and dentistry for the diagnosis or treatment [1, 2]. Bone age, which is routinely requested by pediatricians, endocrinologists and orthodontists, is more accurate in determining the maturation of an individual [3]. In the field of orthodontics, apart from selecting the appropriate appliance to produce the required change in the rate and direction of jaw growth, the treatment timing is critical [1, 4].

Traditionally, analyzing the pattern of ossification of the non-dominant wrist bones using plain wrist radiographs is a fairly predictable method for skeletal age assessment [2]. Hand-wrist radiographs have been the gold standard for determining skeletal age due to simplicity, minimum radiation exposure, and the availability of multiple ossification centers [5]. However, the methods are criticized for the time spent and experience required, inter- and intra-rater variability [6].

Evaluating cervical vertebral maturation (CVM) -as a method to determine skeletal age- can be performed on the lateral cephalometric radiographs using the changes in the size of vertebral bodies as well as shapes of lower and upper borders of C2, C3 and C4 vertebrae [7]. Cephalometry is widely used in orthodontics for the diagnosis, planning, growth and development evaluation and follow up of an orthodontic treatment or progress of a developmental disorder [8, 9]. Thus, in orthodontics, an obvious advantage of CVM evaluation is prevention of additional exposure to radiation by eliminating the need for a hand-wrist radiograph [4].

Baccetti et al. introduced six stages using the morphological changes in the C2, C3 and C4 vertebral bodies, which are commonly observable on a single lateral cephalogram, independent of patient gender [4]. According to this evidence, CVM stages 1 and 2 have been referred to as prepubertal, CVM stage 3 has been referred to as circumpubertal and CVM stages 4, 5, and 6 have been defined as postpubertal [10].

Several studies have stated that CVM is a reliable method of age assessment that can replace hand-wrist radiographs [5, 7]. It has been demonstrated that CVM stages are useful clinical tools to evaluate growth height and mandibular velocities according to the correlation between CVM, chronological age and hand-wrist maturation [11, 12]. However, others have reported that this technique is inherently subjective and influenced by the practitioner’s experience therefore, requires support by other biological indicators [13]. Moreover, some authors believe that due to the high-level of radiographic noise and intrinsic limitations of 2D lateral cephalograms that affect the magnification and image accuracy, the estimation of bone age using CVM may be difficult for practitioners lacking adequate knowledge and experience [4, 13].

Based on the limitations listed above and the fact that accurate image analysis plays a crucial role in achieving a successful orthodontic outcome, automatizing the task will provide time saving, efficiency, accuracy and repeatability in orthodontic treatment planning and assist clinicians in alleviating their enormous workload [4].

Machine learning (ML), uses algorithms to predict the unseen data based on the learnings obtained from intrinsic statistical patterns and structures in data [14, 15]. Deep learning (DL) refers to network architectures with more than one hidden layer that are capable of analyzing complex data structures such as images [14, 16]. DL models require less expert knowledge compared to classical ML methods as they can learn features that adapt to the input data [15].

Recently, the introduction of convolutional neural networks (CNN) algorithms using DL, allows for direct interference, recognition and classification of medical images [17]. CNN has been utilized in various aspects of science including speech recognition, detecting objects, analyzing emotions and face recognition. However, its great breakthroughs in major image competitions have made it a popular technique for medical image analysis and computer visual tasks [18]. In the field of dentistry, CNNs have performed tasks such as caries, bone loss and apical lesions detection as well as classifying, segmenting and detecting anatomic hard- and soft-tissue landmarks [19].

Several AI techniques have been employed for cephalometric radiograph analysis with the focus on auto-identification of landmarks [20]. However, studies on the assessment of skeletal age using lateral cephalograms are in the beginning stage [16]. In addition, CVM analysis on lateral cephalometric radiographs using more recent DL models vary in classification accuracy due to the differences in preprocessing methods and the applied models [21].

Recently, a new imaging technology-cone beam computed tomography (CBCT) is becoming exceedingly popular in the field of orthodontics, which helps in eliminating the problems caused by magnification [22]. It allows orthodontists to evaluate patients’ hard and soft tissue in three dimensions (3D) [23]. Besides, it is superior to conventional CTs due to the lower radiation dose, clearer images, more precision and relatively low cost [5, 24].

Given the importance of CVMs classification in clinical application is to determine the optimum timing for growth modification treatments, and as there is no data available regarding the performance of CNN models to estimate the CVM on 3D radiographs, the objective of the proposed study is to demonstrate the application of CNN in dental imaging for classifying prepubertal, circumpubertal, and postpubertal phases of growth that works in a fully automatic manner without the need for segmentation or annotating (labelling) the images. As the major clinical application of the CVMs classification is to determine the optimum timing for growth modification treatments, we are using this type of categorization which would be more beneficial for clinical decision-making.

## Results

Figure 1 represents a summary of the process from extracting patients’ CBCT images to classifying the phase of growth through CNN models.

**Fig 1.**
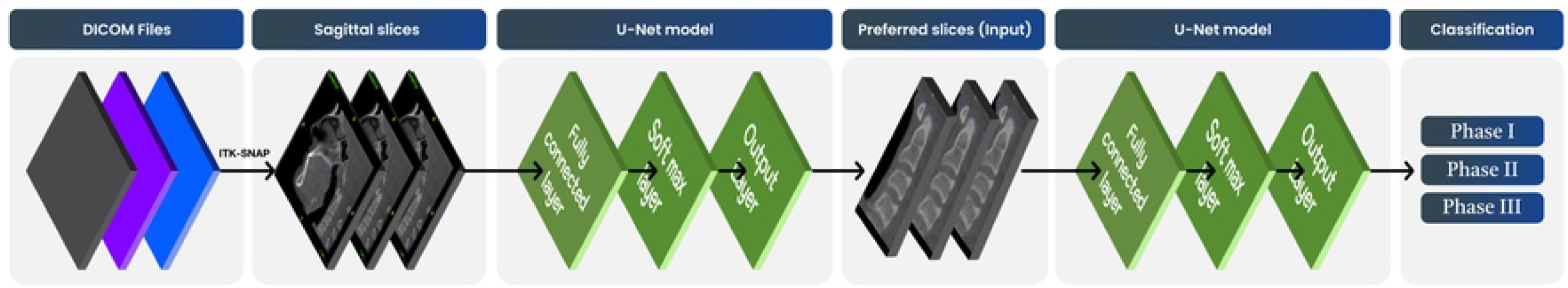
Diagram of the whole process.

Table 1 summarizes the descriptive characteristics of the images and growth phases included in the study. CBCT images belonging to 56 patients (consisting of 536 slices per patient) were first categorized into three growth phases by two orthodontists with an inter-rater reliability of 73%. In cases of conflict, the growth phase was determined by the third orthodontist.

**Table 1.** Descriptive information of the included images.

| Growth phase | Number of patients | Age | Number of slices |
| --- | --- | --- | --- |
| | n (%) | (mean ± SD) | n (%) |
| I | 18(32%) | 8 years and 9 month ± 1 year and 5 months | 536(31.4%) |
| II | 15(27%) | 11 years ± 9 months | 527(49%) |
| III | 23(41%) | 13 years and 7 months ± 1 year and 3 months | 642(37.6%) |

Table 2 demonstrates the performance of the first CNN model to predict preferred vs. nonpreferred views of C2-C4 vertebrae (ROI) on a new set of images as the test dataset. The training and validation accuracies were found to be 91.78% and 88.19%, respectively. According to the table, all slices of new test images including a good vision of vertebrae for classification (n=41) could be predicted correctly.

**Table 2.** Model performance of detecting ROI on the test dataset.

|  |  | Predicted ROI<sup>a</sup> |  |
| --- | --- | --- | --- |
|  |  | Not Preferred | Preferred |
| Actual(true)<br>ROI | Not Preferred | 103 | 72 |
|  | Preferred | 0 | 41 |

<sup>a</sup> ROI: region of interest

Accuracy, recall, precision, and F1-score were calculated using multi-class classification metrics for the second CNN network. Table 3 demonstrates the multi-class classification metrics applied to the validation dataset and a group of 88 images as the testing dataset. The overall accuracy on this set of new slices was found to be 84%. The average classification accuracy of our CNN-based deep learning model was 98.92% and 95.79% on the training and validation datasets, respectively.

**Table 3.**
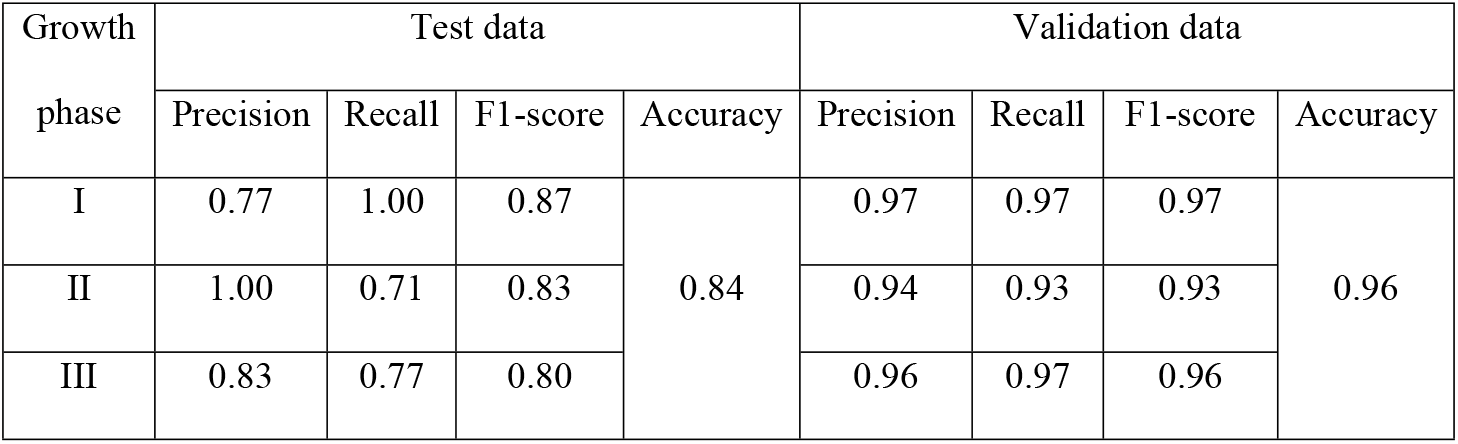
Model performance using the multi-class classification metrics on validation and test datasets for categorizing slices into three growth phases.

| Growth<br>phase | Test data |  |  |  | Validation data |  |  |  |
| --- | --- | --- | --- | --- | --- | --- | --- | --- |
|  | Precision | Recall | F1-score | Accuracy | Precision | Recall | F1-score | Accuracy |
| I | 0.77 | 1.00 | 0.87 | 0.84 | 0.97 | 0.97 | 0.97 | 0.96 |
| II | 1.00 | 0.71 | 0.83 |  | 0.94 | 0.93 | 0.93 |  |
| III | 0.83 | 0.77 | 0.80 |  | 0.96 | 0.97 | 0.96 |  |

Figure 2 also represents the model performance for classifying the cervical vertebrae into 3 growth phases on validation and testing datasets.

**Fig 2.**
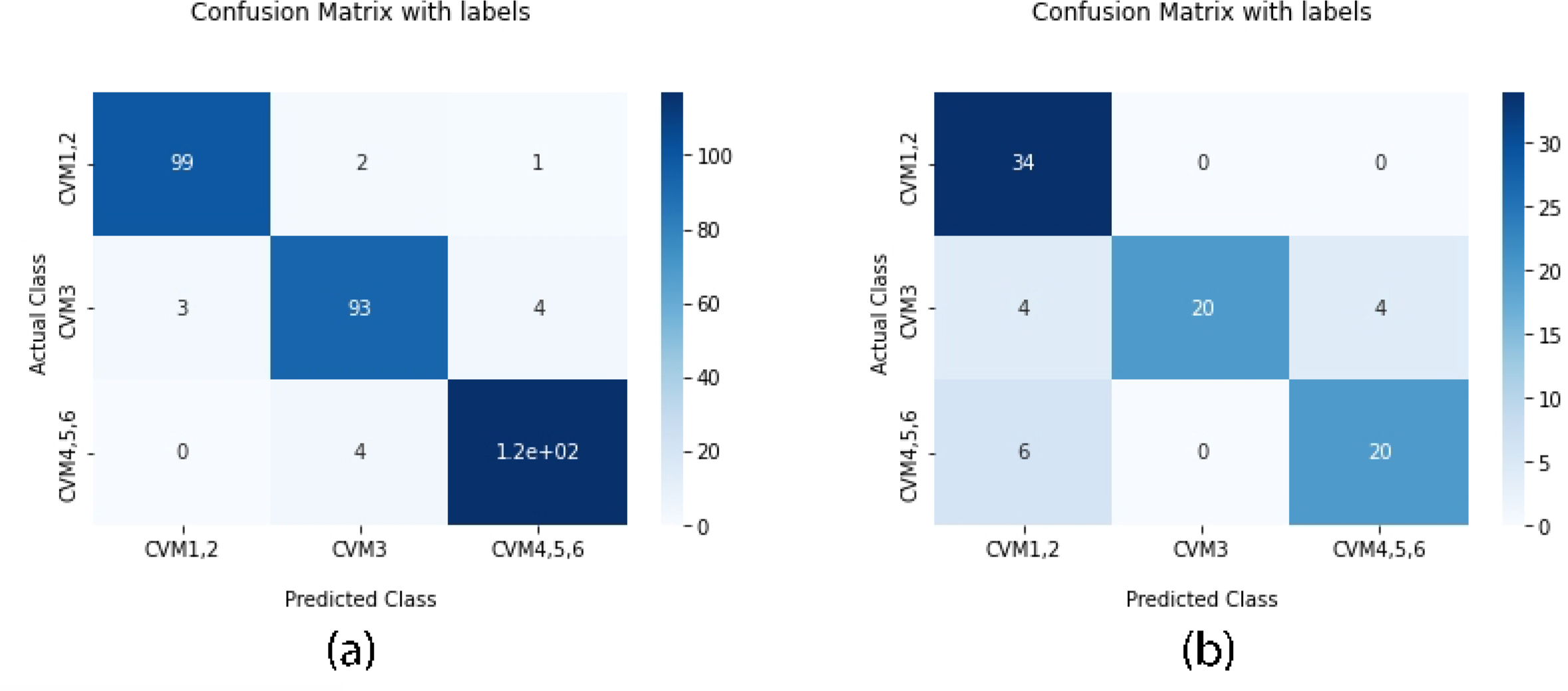
Confusion matrices representing the performance of the model to classify the vertebrae into 3 growth phases on a) validation and b) testing datasets.

## Discussion

In this study, CNN models were designed to classify images according to the presence or absence of the ROI, and then according to the features of vertebrae into three phases of growth. The annotating step was skipped in the proposed model, which resulted in a more time-efficient image pre-processing. To fully automate the process of CVM classification, a recent study by Atici et al. [25] was conducted. They proposed an innovative custom-designed deep CNN to detect and classify the CVM stages. A layer of tunable directional filters was applied to fully automate the procedure and they achieved a validation accuracy of 84.63% in CVM stage classification using 1018 cephalometric images from 56 patients. They stated that this level of accuracy was higher compared to other DL models investigated. Our proposed fully-automated model was successful in determining the growth phase of patients using the CVM staging with a validation accuracy of 95.79%, which is higher compared to Atici et al. findings. This can be due to the higher resolution and accuracy of the input images (CBCT slices) in our study that enhances the training accuracy of the model.

Depending on the task to be performed, various architectures of CNN models have been proposed so far. For instance, Makaremi et al. utilized a semi-automatic CNN-based model to assess the maturation of cervical vertebrae; however, it needed manual segmentation of the region of interest [26]. Since then, many novel methods of image segmentation (such as U-Net) based on FCN have been utilized for medical image analysis [27, 28]. In a study conducted by Seo et al. the performance of six CNN-based DL models were evaluated and compared for CVM analysis on conventional 2D cephalometric images. Inception-ResNet-v2 demonstrated the highest classification accuracy due to its capability of focusing on all three vertebrae (C2-C4) compared to other DL models. They stated that most studied DL techniques classify CVM by focusing on a specific area (region of interest) of the cervical vertebrae. Thus, they suggested that application of high-quality input data and better-performing CNN architectures that are capable of segmenting images will help in creating models with higher performance [29].

Our study used CBCT slices of the vertebrae to determine the skeletal age of the patients. CBCT accuracy and reliability in several aspects of dentistry such as assessment of tumor lesions, orthognathic surgery planning and implant placement have been reported [30]. There is universal agreement that CBCT images are more accurate compared to 2D cephalometrics for craniofacial studies [31, 32]. This can be an explanation for the higher amount of accuracy our model achieved. A recent systematic review by Rossini et al. also showed that 3D cephalometric analysis outperforms the conventional 2D cephalometrics in terms of accuracy and reproducibility [22].

However, the amount of radiation exposure, which is higher in comparison to a 2D cephalogram, is the biggest controversy about its use in dental imaging [33]. It is suggested that CBCT images can be a valid and useful tool for assessment of skeletal age using CVM, although they should not be used solely for that purpose [34].

Our model accuracy on predicting a group of unseen images was greater than 80% with the highest performance at phase I (F1 score:87%), which is consistent with previous studies. According to the literature, CVM stages are sometimes difficult to differentiate according to the continuous nature of morphological changes in cervical vertebrae (McNamara and Franchi 2018). Thus, CS 1 (meaning no development) and CS 6 (maturity) stages are easier to identify. Our model performed well in predicting the CS3 (phase II) with the F1 score of 85%. This was in contrast with a study conducted by Zhou et al. [36] who reported an F1 score of 31% for diagnosing the pubertal spurt on cephalometric radiograph. As the authors mentioned, this could be due to their insufficient training set of CS3 for growth spurt is short and difficult to find in clinical practice.

Hand-wrist radiographs were not used which can be described as a limitation of this study. However, this study focused solely on classifying the patients at their pre-, circum-, and postpubertal growth stages using sagittal slices of the CBCT images and evaluation of the reliability of this method was not taken into consideration.

In contrast to previous studies, we only classified patients according to the three growth phases. However, according to the main clinical application of CVM staging, which is to determine the growth potential of the patients, our classification method can be justified in terms of orthodontic treatment planning and correction of the jaw discrepancies.

In conclusion, our proposed model could automatically detect the ROI (C2-C4) required for CVM staging and accurately classify images into 3 growth phases without the need for annotating the shape and configuration of vertebrae. This will result in developing a fully automatic and less complex system with reasonable performance, comparable to expert practitioners. Classical methods are time-consuming and prone to inter- and intra-rater variability thus, using methods that automate this process will be of value. Expansion and application of utilizing such DL models in clinical practice will enable practitioners to make more accurate diagnosis and treatment planning in a time-saving manner. Moreover, using 3D cephalometric radiographs –which is the primary distinction of the proposed study from the previous ones-could enhance the performance and secure the reliability of the DL model in CVM classification.

## Materials and Methods

This study was approved by the Health Research Ethics Board (HREB) of the University of Alberta (Approval number: Pro00118171). All patients aged between 7-16 years, who underwent CBCT (120 kVp, 5 mA and 4 sec) sagittal views of craniofacial structures between 2013 and 2020 at the University of Alberta, Orthodontics clinic, were included in this study.

The inclusion criteria was as follows:

1. Patients without congenital or acquired malformation of the cervical vertebrae
2. Radiographs with good vision of C2, C3 and C4 vertebrae

Chronological age was collected and calculated based on the date of filming and date of birth. Images of 56 patients were studied. All collected images were kept as DICOM format, so to prepare them for further processing, they were all transformed to PNG images using the ITK-SNAP software (726 * 644 pixels). The sagittal views (cephalometric views), which consisted of 536 slices for each patient studied and classified by two expert Orthodontist Scientists (A. S. and N. A.) with more than 6 years of experience. In the case of any conflicts, a third orthodontist (S. F.) evaluated the slices for determining the class of CVM. CVM was classified into six stages (CS1-CS6) according to the methodology from previous studies [4]. Then slices were grouped into 3 growth phases (I, II, and III) by combining the CS1 and 2 as phase I, CS 3 as phase II, and CS4, 5, and 6 as phase III. Then, the slices were exported into Google Colaboratory for CNN training. Using the original image for classification may lead to poor performance of CNN models since they will classify the cervical stage based on parts other than the shapes of the C2-C4 vertebrae. To overcome this problem, segmentation of the ROI around C2-C4 will enable the classifier model to focus more on these cervical vertebrae [16]. For this purpose, regions of interest (ROI) which included the C2-C4 vertebrae were cropped from the original slices for CVM classification. The result was a collection of 536 slices for each patient (a total of 30,016 slices).

To fully automate the analysis from landmark detection to CVM classification without the need to label the target structures (C2-C4 vertebrae), two classification models capable of classifying the preferable view of C2-C4 vertebrae and estimating the growth phases using a 3D lateral cephalogram were created. In the first model, the resized and cropped ROI obtained from the original image, was used as input for the classifier without segmentation. The classifier model received a fixed image of size 344*350 pixels that fitted the model as an input and classified the image based on presence or absence of the preferable view of all three vertebrae that is required for CVM classification. The output -slices including the preferred vision of vertebrae-was fed to the second CNN model, which predicted the three phases of growth as output. CNNs are types of DL methods consisting of minimum of three layers: input, hidden and output layers [37]. They apply supervised learning technique and called “backpropogation” and have been utilized for various image analysis tasks such as classification, segmentation and landmark detection [38]. In addition to requiring little preprocessing techniques, CNNs are devoid of manual feature handcrafting [3]. The main constituents of a CNN model are: 1) Convolutional layers (the first step) with the purpose of extracting features such as gradients or edges from the input image using the mathematical transformations, 2) Non-linear activation functions, which is sandwiched between any two layers and guides the input signals into output signals required for the NN to act, 3) Pooling layer, which reduces the number of parameters to learn and the amount of computation to summarize the features generated by the convolution layer, and 4) Fully connected layers that are responsible for the interpretation of the feature representations learned by preceding layers [39].

To train the first CNN model for classifying the preferred vs. not preferred views of vertebrae, a labeled dataset is essential. We used 638 slices belonging to two categories from which 127 slices (%20) were selected as validation dataset and remaining slices were used for training. U-Net, a CNN model capable of performing image classification based on fully convolutional networks (FCN) was used [40]. It is a U-shaped model consisting of a contracting path, which goes down to the symmetry point and an expanding path that goes up from that point. The first path, which contains repeated applications of 3-3 convolutions with a rectified linear unit (ReLU) activation, and a 2-2 max pooling operation for downsampling, captures the characteristics of the input image and reduces its size. The second and third path, expanded the image for accurate segmentation and consisted of 3-3 convolutions with a ReLU activation function. The final layer included a 1-1 convolution and the model was compiled using the Adam optimizer and sparse categorical cross entropy loss function. The final output was a collection of a range of 21-35 slices (28.17± 3.06) for each patient thus, a total number of 1705 slices from 56 patients (536 slices for phase I, 527 for phase II, and 642 slices for phase III) were finally obtained. From each phase, a collection of slices belonging to a patient was randomly selected as the test dataset, thus 88 slices (34, 26, and 28 slices representative of growth phase I, II, and III, respectively) were not input the second model

The second CNN model to classify the slices into three growth phases was trained using 1617 slices (out of 1705 total slices), which were split into training (1294 or 80%) and validation (323 or 20%) datasets. The architecture of the second model was the same as the first one except for removing the dropout from the third hidden layer, and the number of epochs (25 vs. 3). After training the model, 88 unused slices were used as the testing dataset to evaluate the performance of the model using multi-class classification metrics. All calculations and computations were completed using python (TensorFlow, NumPy, Matplotlib, and Keras packages).

## Data Availability

Code is available upon request to corresponding author. Images are not available due to privacy.

## Acknowledgement

The authors would like to thank Ashley Fossen for her assistance in data anonymization and for providing patients’ DICOM files.

